# Alzheimer’s Disease Assessments Optimised for Diagnostic Accuracy and Administration Time

**DOI:** 10.1101/2021.07.01.21259858

**Authors:** Niamh McCombe, Xuemei Ding, Girijesh Prasad, Paddy Gillespie, David P. Finn, Stephen Todd, Paula L. McClean, KongFatt Wong-Lin, Alzheimer’s Disease Neuroimaging Initiative

## Abstract

**Objective:** Despite the potential of machine learning techniques to improve dementia diagnostic processes, research outcomes are often not readily translated to or adopted in clinical practice. Importantly, the time taken to administer diagnostic assessment has yet to be taken into account in feature-selection based optimisation for dementia diagnosis. We address these issues by considering the impact of assessment time as a practical constraint for feature selection of cognitive and functional assessments in Alzheimer’s disease diagnosis.

**Methods:** We use three different feature selection algorithms to select informative subsets of dementia assessment items from a large open-source dementia dataset. We use cost-sensitive feature selection to optimise our feature selection results for assessment time as well as diagnostic accuracy. To encourage clinical adoption and further evaluation of our proposed accuracy-vs-cost optimisation algorithms, we also implement a sandbox-like toolbox with graphical user interface to evaluate user-chosen subsets of assessment items.

**Results:** We find that there are subsets of accuracy-cost optimised assessment items that can perform better in terms of diagnostic accuracy and/or total assessment time than most other standard assessments.

**Discussion:** Overall, our analysis and accompanying sandbox tool can facilitate clinical users and other stakeholders to apply their own domain knowledge to analyse and decide which dementia diagnostic assessment items are useful, and aid the redesigning of dementia diagnostic assessments. Clinical Impact (Clinical Research): By optimising diagnostic accuracy and assessment time, we redesign predictive and efficient dementia diagnostic assessments and develop a sandbox interface to facilitate evaluation and testing by clinicians and non-specialists.

## I. Introduction

**D**ementia, of which Alzheimer’s disease (AD) is the most common type, causes enormous global socioeconomic burden [1]. Adding to the urgency of the problem due to ageing societies, is the sub-optimal dementia care pathway, that impacts everything from diagnosis to management of care (e.g. [2], [3]). For people with dementia to receive appropriate treatment and support, careful assessment for diagnosing dementia is necessary. There are several types of clinical assessments and associated markers for dementia, from clinical history, biological (e.g. blood- or brain-based) assessments, to cognitive and functional assessments (CFAs) [4]. CFAs provide information on dementia symptoms and are highly accurate diagnostic markers [5], forming a key component of the clinical care pathway [6]. Although they vary in terms of diagnostic accuracy, sensitivity and specificity [6], and are not always administered in a standardised way across clinical practices [4], they perhaps form the most critical component in dementia diagnosis [4].

Many studies on dementia treatment pathways have ascertained that the societal burden of dementia can be partially reduced by early diagnosis, particularly from a reduction in the potential cost of care home treatment [7]. Further, early diagnosis and intervention can increase quality of life for people with dementia and their carers [8]. Moreover, a meta-analysis on dementia diagnosis in primary care suggests that fewer than half of dementia patients were accurately diagnosed [9]. Critically, in addition to the difficulty of diagnosing dementia, clinicians’ consultation time is also typically limited [4]. In particular, general practitioners usually only have about 10-15 minutes for a consultation [10], [11].

To deal with the above issues, recent studies have explored composite (assessment) scales as tools for dementia assessments (e.g. [12], [13]). Specifically, a composite test assesses different domains of cognition or everyday life functioning, and then combines over the scores for each domain to yield an overall score. Many of these composite assessments are mere combinations of previously developed assessment scales, which could be unwieldy or potentially duplicate information [12]. Importantly, it is unclear whether composite assessments can actually perform better (e.g. higher sensitivity and specificity) than the current battery of assessments used in clinical practice. Moreover, these studies often did not consider large sets of assessments, which may require automation in the selection process, e.g. using machine learning algorithms.

The use of machine learning approaches in dementia research has been facilitated by openly available dementia datasets [4]. They often include a wide range of dementia assessments, including CFAs, with large sample size, thus providing a rich data source to develop and apply machine learning techniques towards improving dementia diagnosis and prognosis [14], [15]. In particular, with the large number of variables in these open datasets, and in clinical datasets, a frequently employed machine learning approach is feature selection [16], which seeks to identify which variables (features) are relatively more useful for building a computational (e.g. predictive) model. The resultant computational model can then guide effective clinical diagnosis (e.g. identifying level or class of disease severity using only a small subset of data features). Thus, this makes them appropriate for use in clinical decision support systems [4].

Feature selection methods can be classified as either univariate methods, which evaluate individual features without considering feature dependencies, or multivariate methods, which evaluate patterns in subsets of features [16], [17]. Feature selection techniques are also grouped as filter techniques (independent of the classifier algorithm), wrapper techniques (interact with the classifier to find the features which are most useful for classification), and embedded techniques (some form of feature selection is incorporated within the training of the classifier algorithm) [16], [17]. An emerging field in the realm of feature selection for medical data is cost-sensitive feature selection methods, which penalise ‘costly’ features (e.g. time cost to administer an assessment) in the evaluation criteria [16].

Feature selection could be used for modelling prognosis of dementia. For example, a study that used cognitive assessment items to build a machine learning prognostic model could predict progression from mild cognitive impairment (MCI, frequently a prodromal stage of dementia) to dementia in both 3-year and 5-year time windows [18]. Another dementia progression study had performed meta-analysis across multiple studies that use CFAs to classify MCI and predict MCI-to-dementia progression, and found them to perform better for diagnostic than prognostic predictions [19]. In terms of data pre-processing, issues such as missing data handling and class balancing were addressed, and genetic algorithm was applied to select features predictive of disease status at different stages [20]. In terms of AD diagnosis, a study had trained a deep learning network and used Recursive Feature Elimination feature selection in building a classifier to diagnose AD stage, and found the time orientation features of the Mini Mental State Examination (MMSE) to be the most powerful predictors of the evaluated features [21].

Despite such studies, there are only a few studies focusing on evaluating the finer assessment units or items (i.e. specific questions) for dementia diagnosis. For instance, in terms of applying feature selection algorithms on specific assessment items for AD diagnosis, a study had used false-discovery rate feature selection algorithm combined with expert input from neuropsychologists to select specific assessment items which are diagnostic of AD stage [22]. Another study that evaluated the granular aspects of the cognitive and functional assessments found 4 neuropsychological features which could differentiate between clinically impaired (MCI or dementia) and non-impaired individuals with 94.5% sensitivity and discussed that using these 4 features to diagnose could reduce assessment time to an average 15 minutes testing time plus a 15 minute delay [23].

Despite the above studies, it is still unclear to what extent such feature selection approaches for searching optimal dementia assessments are conducive in clinical practice. Importantly, the cost of dementia assessment time, a practical constraint, has not been considered in the above studies. In fact, some CFAs take much longer to administer than others [6] and thus may not be considered suitable for screening appointments in primary care (more comprehensive assessments are often conducted in secondary care). In fact, assessment or physician time is one of the most critical resources which must be optimised in the context of dementia assessment to relieve pressure within the healthcare service [23]. Hence, to properly translate feature selection techniques to clinical use, diagnostic accuracy versus assessment time trade-off must be considered.

In this paper, we address this issue by applying a cost-sensitive algorithm for feature selection which takes into account the trade-off between classification accuracy of dementia severity and the total time available for dementia diagnostic assessment. In particular, we apply this algorithm to an open dementia dataset to identify CFA features that balance the classification accuracy of dementia severity and the time cost of individual items within each assessment. Further, to encourage clinicians, health economists, policy-makers and other stakeholders to be more involved in the adoption of machine learning based solutions, we develop, on top of this algorithm, a user-friendly graphical user interface (GUI) to act as a cost-based assessment optimisation sandbox-like toolbox for such users to update or test any assessment information using their domain knowledge, to rediscover the most efficient assessment items based on the individual clinician’s available consultation time, and to redesign and streamline dementia diagnostic assessments.

## II. Methods and Procedures

### A. Data

We made use of the Alzheimer’s Disease Neuroimaging Initiative (ADNI) open datasets (adni.loni.usc.edu). In particular, we focus on the CFAs of the data as they form a key component of dementia clinical assessment [6]. Further, previous studies have shown that CFAs, when considered in machine learning approaches, can achieve relatively high accuracy in identifying AD severity [22], [5], [24], [21], [20], [23].

Specifically, the considered CFAs were MMSE [3], Montreal Cognitive Assessment (MoCA) [25], Alzheimer’s Disease Assessment Scale (ADAS) [2], Functional Activities Questionnaire (FAQ) [26], Everyday Cognition – Patient scale [27], Geriatric Depression Scale [28], and Neuropsychological Battery (NB). Within these CFAs, 113 assessment items were combined with patient demographic variables (age and education level) into one data table for feature selection. Although the ADNI data is longitudinal, we retained CFA data from participants’ first visits only to avoid potential confounding effects such as those due to practice effects [29]. Further, assessment items that cannot be assessed separately were combined with each other. For instance, the MMSE item `Spell WORLD backwards`, where each letter is scored separately in the data, was combined into a single item labelled as `mmse_MMWORLD`. Detailed information of all included assessment items is shown in Supplementary Table 1.

We used Clinical Dementia Rating Sum-of-Boxes (CDR-SB) rating as the objective measure of dementia severity due to its ability to track both cognitive and functional disability in AD and dementia stages [30], [31], [32], and our previous work that shows strong correlation with clinical diagnosis [5] and practical use for clinical decision support system [24]. CDR-SB was re-coded into 5 classes of Alzheimer’s disease (AD) severity as described in [31]. As there were very few cases of moderate or severe AD in the dataset, all AD subcategories were amalgamated into one category, creating 3 groups: Cognitively Normal (CN); Mild Cognitive Impairment (MCI); and Alzheimer’s Disease (AD), incorporating the mild, moderate and severe AD classes.

### B. Computational Methods

#### 1) Feature Selection for the most diagnostically valuable assessment items

Three different feature selection methods were used. The feature selection methods were chosen because they are multivariate (i.e. they evaluate each feature in the context of other features rather than ranking individual features) and their approaches of evaluation are very different. Specifically they were: (i) Correlation-Based Feature Selection (CFS) [33], [34], which evaluates the worth of every possible subset of features by considering the individual predictive ability of each feature along with the degree of redundancy between them (see Supplementary Algorithm 1); (ii) Boruta [35], a wrapper-based feature selection algorithm based around a Random Forest (RF) classifier, which evaluates all features to see if they perform better than randomised “shadow features” when the classifier is applied (see Supplementary Algorithm 2); and (iii) Consistency [36], [34], which selects features based on whether they are consistent - i.e. whether the same feature values consistently co-occur with class label (see Supplementary Algorithm 3). Each algorithm was run on five different folds of the data (five-fold cross-validation), creating five sets of items selected by each algorithm (Fig. 1).

**Fig. 1.**
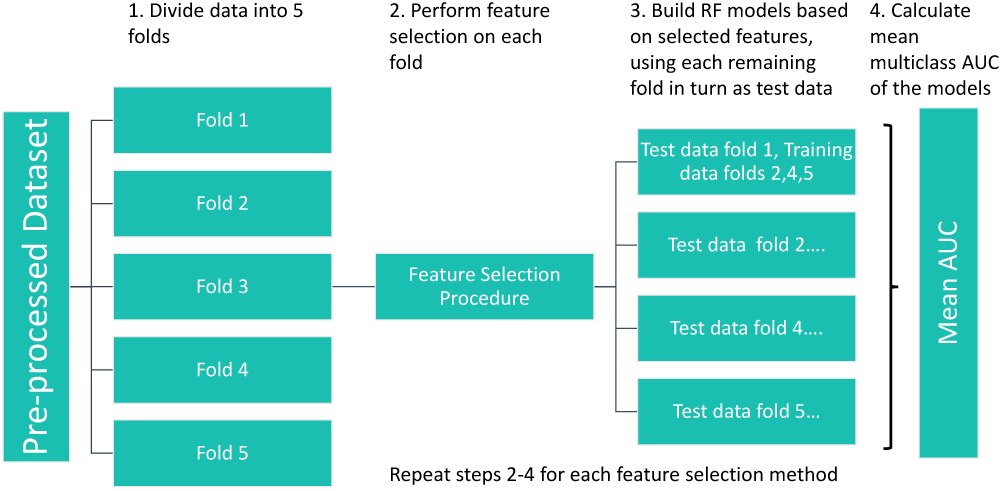
Pre-processing and analytical pipeline for selecting accuracy-optimised sets of dementia assessment items. Note: Step 2 shows an instance of feature selection - performed on Fold 3.

As previous work [37] testing multiple classification algorithms has shown the RF classifier to be an effective classifier on ADNI data, and also more robust to different data pre-processing methods than other classifiers [38], we built RF models using the RandomForest package in R [39] to predict CDR-SB using each of the assessment item sets selected by the above three feature selection algorithms. The fold of the data which had been used for feature selection was excluded from the training and testing datasets, with each of the remaining folds of data in turn used as the test dataset, while the remaining three folds are used as the training dataset (Fig. 1). The multiclass receiver operating characteristics area-under-curve (AUC) of these models was averaged over the four folds for each of the five sets of selected features. Multiclass AUC was calculated by the process defined in [40] using the pROC package [41]. This method of calculating multiclass AUC extends the binary AUC concept to multiple classes by calculating the pairwise AUCs of each class against every other class and then averaging the results.

#### 2) Accuracy-cost optimisation

We use multiclass AUC, as defined in the section above, as our diagnostic accuracy metric in this section and the Results section. The costs being optimised are the assessment times for the individual diagnostic assessment items. We used known overall assessment times for CFAs in clinical practice to estimate an assessment time for each assessment item that was selected more than once - i.e. in multiple folds and/or by multiple algorithms - by our feature selection algorithms. These items are shown in Supplementary Table 2. In most cases, to assign a time cost to an assessment item, the total time for the whole assessment (e.g. MMSE) as given in the literature [3], [25], [2], [26], [27], [28] was divided by the total number of items in that assessment. We used the ADNI procedures manual and the comprehensive information in [42] to augment the process. (Supplementary Table 2 provides further details on the time-cost estimation process.) These values were then used as the default values in the GUI tool (see Section III-C) where they can be amended by the GUI user, depending on e.g. the user’s experience of conducting the assessments in practice.

We modified the code for the CFS algorithm [33] from the FSelector package [34] in R, to use the cost-sensitive feature selection framework [43]. This modified version of CFS (cost-sensitive CFS) incorporates a cost weighting parameter, *λ*, which can be varied to reflect different values of cost weighting for the features. If *λ* is set to zero, the algorithm performs like a typical CFS without time cost. We used CFS here as its evaluation function (a mathematical formula which is applied to every feature set to determine its usefulness) can be easily modified to incorporate time costs, unlike Boruta which does not use an evaluation function (see Supplementary Algorithms 1 and 2), and the features sets selected by CFS performed better than the feature sets selected with the Consistency algorithm (see Fig. 4).

**Fig. 2.**
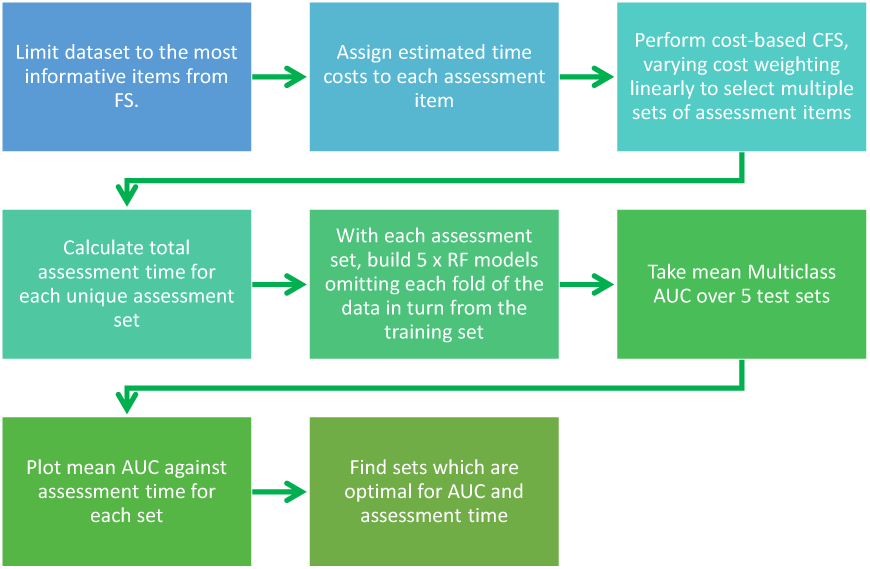
Analytical pipeline for optimising accuracy-cost dementia assessment items.

**Fig. 3.**
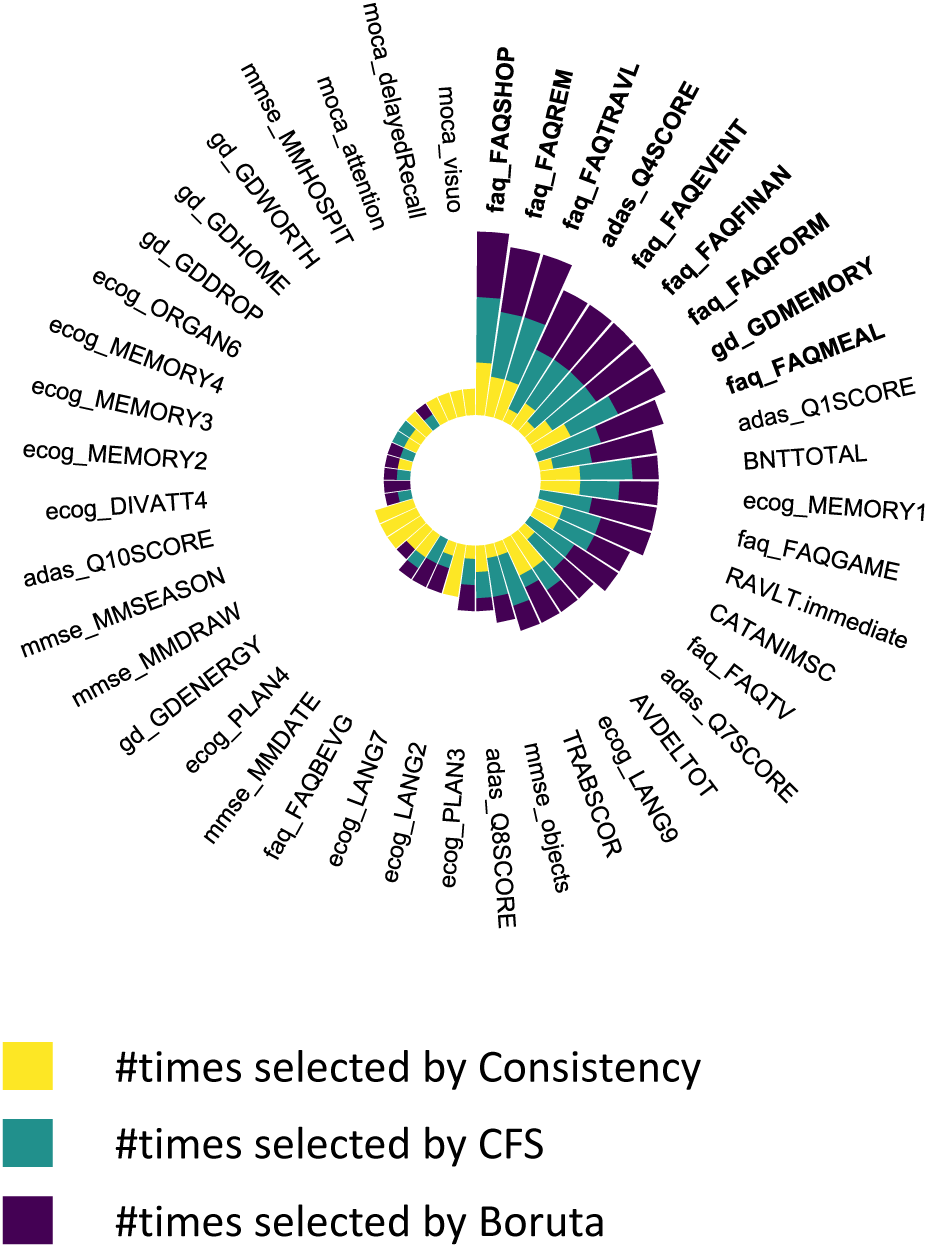
The most consistently selected CFA items. Only data features selected more than once in the 5× 3 iterations feature selection process are shown. Bold text: Features consistently selected 10 or more times.

Specifically, the CFS algorithm finds an optimal set of features which correlate with the class variable and do not correlate with each other. It uses a “*Merit*” heuristic to evaluate the best feature set, described by

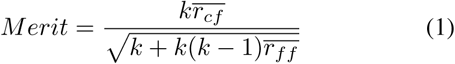

where *k* is the number of features in the set, 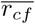 is the average feature-class correlation, and 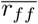 is the average feature-feature correlation. In the implementation in the FSelector package [34], correlation between discrete features is measured by mutual information, and for continuous features the correlation coefficient is used. It can be seen that the merit heuristic incorporates a penalty to favour smaller sets of features over larger sets. The best first search is used to find the subset with the highest ‘merit’. It should be noted that since the best first search is not exhaustive, a different randomisation may lead to travelling down a different search path, and therefore the results from CFS will not necessarily be identical every time the algorithm is applied to the same dataset.

The cost-based CFS extends the above function by adding a cost penalty to the evaluation function [43]

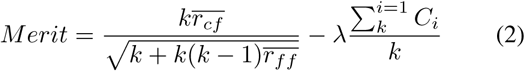

where *C*_*i*_ is the feature cost of item *i*, which in our case is the time of each assessment item, and *λ* is the cost weighting parameter which can be varied to increase or decrease the importance of feature cost in the merit heuristic.

We restricted the columns of the dataset to the features selected more than once by feature selection methods and applied cost-sensitive CFS to this data. We varied the *λ* parameter from 0 to 0.05 and recorded the set of assessment items selected at each point of variation, along with the total estimated time for the selected set of items. The appropriate range of values for the *λ* parameter will be different depending on the values of the feature costs. For the case of *λ* having zero value, the algorithm is identical to CFS. Values of *λ* greater than 0.01 did not generate additional sets of features for analysis.

To predict CDR-SB using each of the assessment item sets selected by the cost-sensitive CFS, we again built RF models. Also as before, each of the five folds of data used in Section II-B1 was used in turn as the test dataset, with the remainder of the data used as the training dataset (Fig. 1). The multiclass AUC was calculated and averaged over the five folds as described in Section II-B1. The entire process for the accuracy-cost optimisation is summarised in Fig. 2.

### C. GUI development

We developed a GUI-based sandbox-like toolbox using the accuracy-cost optimisation algorithms described above, which can allow non-technical users to input their own time estimates for any of the 113 assessment items in our dataset, perform cost-sensitive feature selection, and calculate the AUC of an RF model to predict AD severity (CDR-SB value) using any chosen set of assessment items from the base dataset.

We implemented the GUI in RShiny [44] using the DT package [45] in R to create an editable data table, and the shinyWidgets package [46] to build the interface for user selection. For computational efficiency, this GUI interface does not use cross-validation as above, but uses 25% of the underlying data for feature selection, and 75% of the remaining 75% for model training, while the remainder is test data.

### D. Software and hardware

The above analyses and algorithms were run within R Studio version 1.146 on a Windows machine with eight memory cores, Intel i7 processor, 16GB RAM and R version 3.5.2 installed. The codes, including the RShiny code for the GUI, are available at: https://github.com/mac-n/Rshiny-app.

## III. Results

### A. Small Subsets of High-Performing Assessment Items Consistently Selected

There were variations in the mean number of data features selected by each feature selection algorithm; 19 features per fold for CFS, 16 for Consistency, and 19 for Boruta. All the data features selected more than once in the 5*×* 3 iterations (Fig. 1) during feature selection are shown in Fig. 3 and in Supplementary Table 2. Fig. 3 also shows that a small number of features were selected consistently by the algorithms. The features selected 10 or more times are labelled in bold text in Fig. 3; there were 9 of them. Most of the data features do not appear in Fig. 3 because they were selected only once (16 of the features) or never selected (54 of the features). Note that the assessment items from the FAQ assessment were frequently selected by the 3 feature selection algorithms, and this was consistent with our previous work that showed the total FAQ score to be highly accurate for classifying AD severity [5], [24]. The multiclass AUC of each set of features selected is shown in Fig. 4, varying around between 0.86 and 0.94. The AUC of the complete feature set without feature selection is shown by the dashed line in Fig. 4; most of the selected feature subsets outperformed this complete set. Information on the constituent item sets of every item in Fig. 4 has been up-loaded to GitHub at https://github.com/mac-n/Rshiny-app/blob/master/JTEHM_McCombe_data.xlsx.

**Fig. 4.**
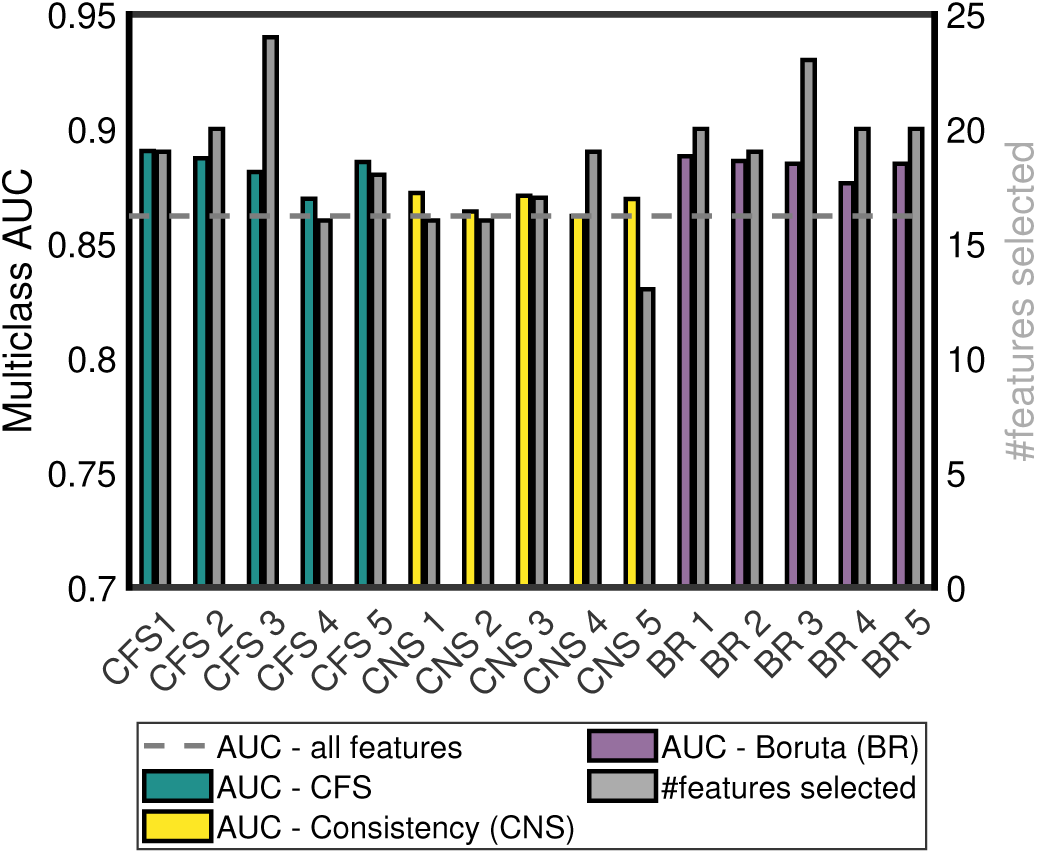
Multiclass AUC of each selected feature set. Dashed line: AUC using all features.

### B. Selected Assessment Item Sets Optimised for Assessment Time and Classifier AUC

For convenience, we made use of the selected features in Fig. 3 for the cost-sensitive feature selection. The plot of total assessment time against classification AUC for different values of the cost weighting parameter *λ* (see Section II-B2) is shown in Fig. 5. Information on the constituent item sets of every item in Fig. 5 has been uploaded to GitHub at https://github.com/mac-n/Rshiny-app/blob/master/JTEHM_McCombe_data.xlsx. It should be noted that when *λ* was above a certain value (0.01), only two items were selected (`mmse_MMDATE` and `mmse_MMHOSPIT`) and the AUC declined rapidly to 0.6. With this single exception, the selected assessment subsets (Fig. 5, top filled circles) were comparable in terms of multiclass AUC values. The limited range of AUC values was expected due to our earlier feature selection process, i.e. these subsets of assessment items had been previously optimised in terms of AUC (see above). In particular, these selected assessment subsets generally had higher AUC values than standard individual (and not optimised) assessments such as MMSE, MoCA, and ADAS (compare filled circles to opened circles in Fig. 5). However the AUC value of FAQ was on par with the optimised assessment subsets, which was consistent with our earlier work supporting FAQ’s high predictive power of AD severity [5], [24].

**Fig. 5.**
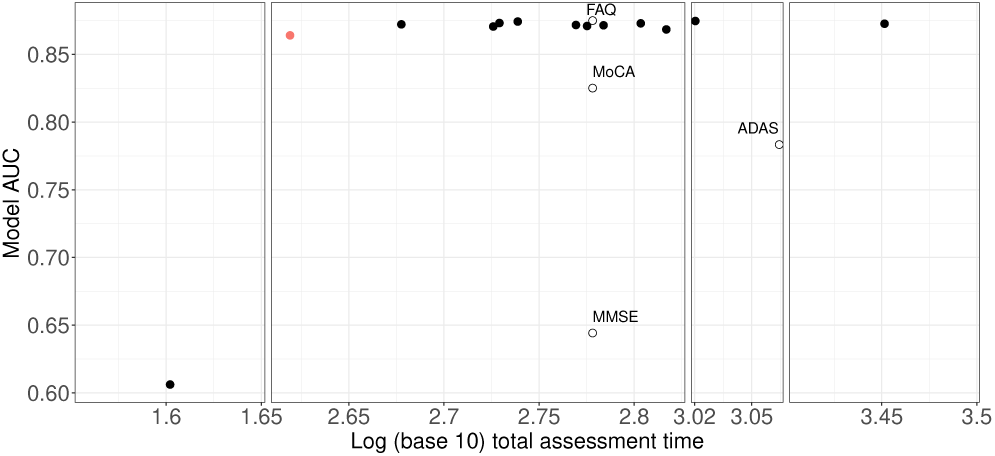
Wide range of (estimated) total assessment times of accuracy-cost optimised feature sets. Vertical axis: AUC values based on 3-class RF classifier. Horizontal axis: Logarithmic scale of (estimated) total assessment time. AUCs of accuracy-cost optimised sets (filled circles) higher than ADAS, MMSE and MoCA complete standardised assessments while on par with FAQ complete assessment (black opened circles).Orange filled circle: Optimal item set with the shortest total assessment time (see Table I for details on its specific items).

In contrast to AUC values, the assessment times of the selected item sets varied much more widely. When *λ* was set to zero (i.e. time cost not being considered in the feature selection process), the total assessment time was found to be relatively long at 2288 seconds, although the AUC value was 0.879 (the highest in Fig. 5). The constituent items of this item set, with associated estimated assessment times, were: `faq_FAQSHOP` (60); `faq_FAQREM` (60); `faq_FAQTRAVL (`(60); `adas_Q4SCORE (`600); `faq_FAQEVENT` (60); `faq_FAQFINAN (`60); `faq_FAQFORM` (60); `gd_GDMEMORY` (28); `faq_FAQMEAL` (60); `RAVLT.immediate` (900); `ecog_MEMORY1` (60)); `adas_Q7SCORE` (200); `faq_FAQGAME` (60); `faq_FAQTV` (60); `mmse_objects` (300); and `adas_Q8SCORE` (200).

The shortest assessment time calculated for any of the viable assessment time sets was 416 seconds. This item set, with an AUC of 0.865, is highlighted in blue in Fig. 5 and its features detailed in Table I.

**TABLE I.** accuracy-cost optimised feature set with the fastest total assessment time.

| Item | Question | Estimated Time (sec) |
| --- | --- | --- |
| faq_FAQSHOP | (Patient's degree of difficulty with) Shopping alone for clothes, household necessities, or groceries | 60 |
| faq_FAQREM | Remembering appointments, family occasions, holidays, medication | 60 |
| faq_FAQTRAVL | Traveling out of the neighborhood, driving, or arranging to take public transportation. | 60 |
| faq_FAQEVENT | Keeping track of current events. | 60 |
| gd_GDDROP | Have you dropped many of your activities and interests? | 28 |
| gd_GDMEMORY | Do you feel you have more problems with memory than most? | 28 |
| mmse_MMDATE | What is today's date?. | 20 |
| mmse_MMSEASON | What season is it? | 20 |
| mmse_MMHOSPIT | What is the name of this hospital (clinic, place)? | 20 |

Although the selected assessment subset had both AUC value and overall assessment time similar to those for FAQ, it should be noted that this optimal assessment subset contained a combination of orthogonal items (due to the nature of our feature selection algorithm) from both cognitive (MMSE) and functional (FAQ) assessments (Table 1) - this would probably be preferred to FAQ alone in clinical practice, as cognitive impairment is a necessary criterion for dementia diagnosis [6].

Taken together, we found that there were subsets of assessment items optimised for diagnostic accuracy and total assessment time which could perform better, in terms of classification AUC and/or assessment time, than several of the individual standard complete (MMSE, MoCA and ADAS) assessments (Fig. 5) and the full set of complete assessments (Fig. 4), while on par with the complete FAQ assessment (Fig. 5) (despite its lack of evaluating the cognitive aspects).

### C. GUI Tool for Further Exploration

The method for estimating assessment item time costs described in Section II-B2 can be refined for future use. For instance, clinicians who work with dementia patients may have access to their own estimates. Further, health economists or policy-makers who are considering the redesigning of dementia diagnostic assessments may experiment with various scenarios for trading off the time cost of assessment with diagnostic accuracy. To extend the impact of our work, we developed an R Shiny based graphical user interface (GUI) sandbox-like tool for exploring cost-sensitive feature selection and diagnostic accuracy on subsets of the CFA assessment items, building on top of the abovementioned algorithms. This software application is made available at https://mac-n.shinyapps.io/costcfs/. A video demonstrating the use of the app is available in the GitHub repository: https://github.com/mac-n/Rshiny-app. A screenshot of the application’s full interface upon opening the application is shown in Supplementary Figure 1. As illustrated, the GUI design is minimalist and does not require computational expertise. The app has been tested on OS X (Mohave version) and Windows 10, on the Safari, Chrome, Firefox and Edge browsers.

Upon opening the application, a selection box (Fig. 6a) allows users to choose any subset of the assessment items in our dataset for analysis. For instance, some clinicians may only use specific assessment items (e.g. specific assessment questions within MMSE and MoCA) due to the nature of their practice. To further assist this, we also created a drop-down menu (Fig. 6b) listing subsets expected to be of interest to users, including: (i) assessment items shown in Fig. 3 which were used for the analysis described in Section II-B2; (ii) complete set of assessment items in MMSE; (iii) complete set of assessment items in MocA; (iv) complete set of assessment items in ADAS; and (v) complete set of assessment items in FAQ. These specific assessments were placed here as MocA and MMSE are often used in clinical practice [6], while ADAS is more frequently used as a benchmark for dementia symptom evaluation in drug trials [6]. Moreover, FAQ was highlighted in the menu since our current work and previous studies (e.g. [5], [24]) have indicated its potential strong diagnostic utility.

**Fig. 6.**
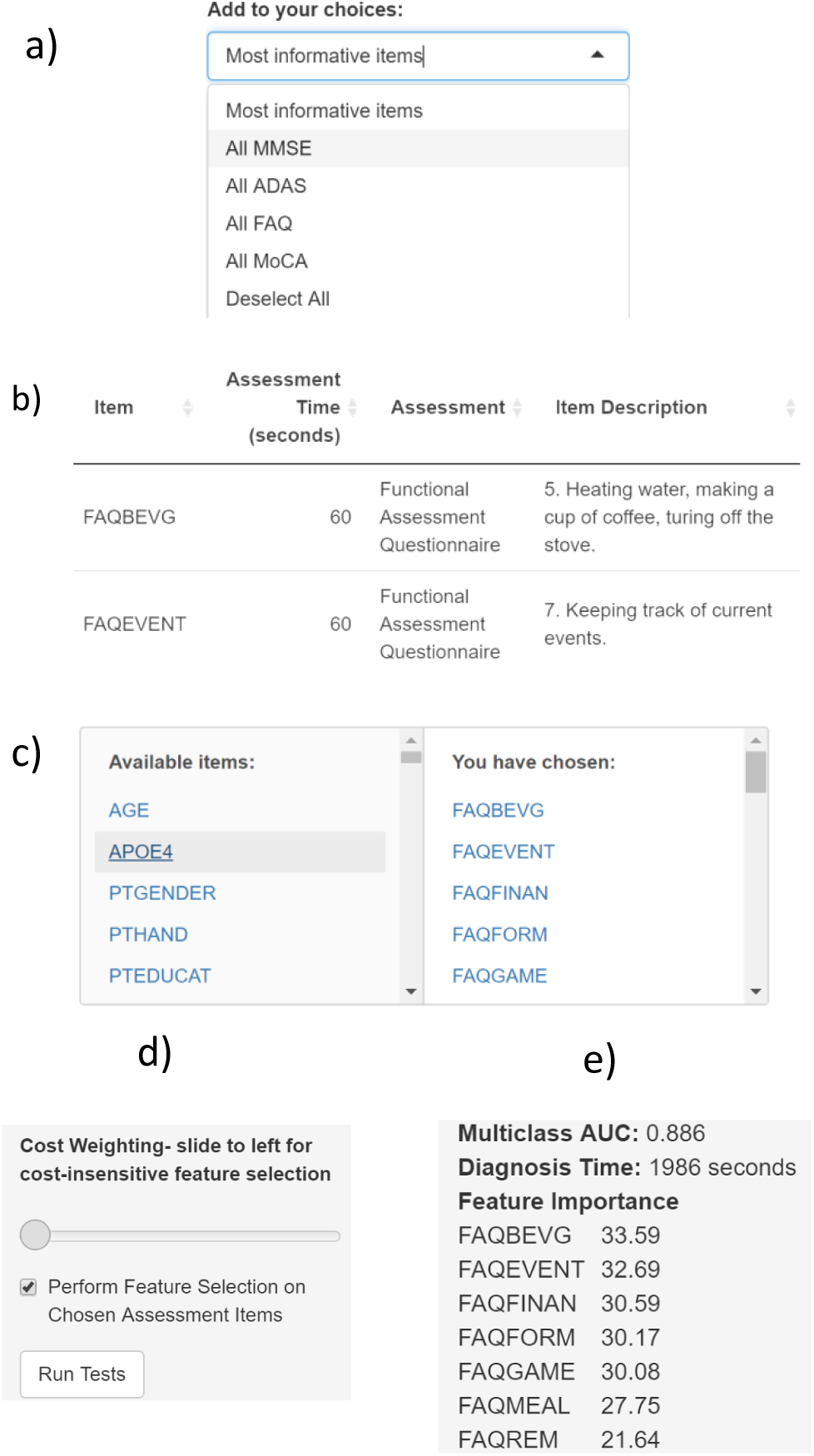
Different features of the cost-benefit analytical sandbox tool. (a)-(e): Order of features appearing during usage.

Upon choosing the subsets of assessments for analysis, the items will be displayed in a table (Fig. 6c). The user may then edit the costs (time, in seconds) associated with these features. Note that we have not estimated time costs for features other than those shown in Fig. 3; other features have been assigned a default value of 1001 seconds, which a user can easily edit.

Cost-sensitive CFS feature selection is toggled on and off with a checkbox (Fig. 6d) and the cost parameter is varied with a slider (Fig. 6d.) If the user wishes to perform feature selection without considering feature costs, the parameter may be set to zero. The algorithm is activated with the “Run Tests” button. (Fig. 6d). If feature selection is toggled on, an RF classifier will be built and tested using the subset of user-chosen assessment items selected by the algorithm. Otherwise, the classifier will be built using all the user-chosen assessment items.

Upon completing the calculation, the classifier for the 3-class classification AUC, the overall assessment time for the assessment items used in the classifier, the names of all the assessment items used and their feature importance value in the model will be displayed (Fig. 6e).

## IV. Discussion and Conclusion

Our cost-sensitive feature selection analysis had led to the identification of combinations of cognitive and functional assessment (CFA) items (see Table I optimised for total assessment time and classification AUC of (CDR-SB based) AD severity. The identified subsets of accuracy-cost optimised assessment items were found to outperform existing assessments. Despite the small sets of assessment items, our 3-class classification AUC values resided within the range of 0.86-0.94, which were rather high. Importantly, most of the identified accuracy-cost optimised assessment sets had total assessment times under the typical clinical consultation time of 15 minutes.

In comparison, previous work had only optimised for the accuracy for classifying AD (e.g. [22], [19]). In particular, a study [22] had used the ADNI dataset with CDR-SB as the outcome variable and found very high sensitivity and specificity for the sub-assessments selected by its false discovery rate feature selection algorithm. Many of the same sub-assessments were selected as in our work, e.g. FAQ items and ADAS Q4, but the previous study [22] also included total scores from assessments such as FAQ, MMSE etc, and total scores frequently emerged as highly predictive in feature selection. In comparison, our work did not include total scores and we optimised for assessment time as well as classification accuracy (AUC) of AD severity.

In another study [21], which used a different dataset (the Seoul Neuropsychological Screening Battery), 46 data features (most of which did not overlap with those in ADNI) and a deep learning classifier were used. It was shown that 3-class classification accuracy of over 90% could be obtained using only 12 of the 46 features. Of the features which did overlap with ADNI, it was found that MMSE orientation to time and MMSE 3-word recall were highly predictive for cognitive impairment classification. Our current study also found these features to be predictive, with the `mmse_objects`, `mmse_MMSEASON` and `mmse_MMDATE` items consistently selected by feature selection methods (Fig. 3) and the `mmse_MMSEASON` and `mmse_MMDATE` items selected by the accuracy-cost optimisation algorithm (Table I.

A separate work [23] had optimised CFA features in the ADNI dataset to detect MCI and found a set of 4 neuropsychological features which could classify MCI with 94.5% sensitivity. Unlike other work discussed here, this work did explicitly discuss assessment time, although they did not computationally optimise for assessment time. Notably, they estimated that their 4 identified features (`delayed WAIS Logical Memory`, `trail-making`, `patient` and `informant memory questions`) would take about 15 minutes of clinician’s time to administer. However, an additional 15-minute delay would entail due to the delayed memory recall task within the `delayed WAIS logical memory` assessment. Such a long delay would likely render this set of assessments to be unsuitable for use in brief clinical consultation routinely available in primary care. Interestingly, their best assessment set included `patient` and `informant memory` questions, and the `trail-making` assessment, which were also highly selected by our algorithms (`gd_GDMEMORY`, `faq_FAQREM` and `TRABSCOR`; see Fig. 3 and Supplementary Table 2). Our final optimised assessment also included the `patient` and `informant memory` questions (`gd_GDMEMORY` and `faq_FAQREM`; see Table I.

Cost-sensitive analysis is an important aspect of feature selection for medical applications [16]. However, in dementia data science research, such analysis is often not taken into consideration. Hence, this leads to many machine learning based solutions not reaching their fuller impacts, and not readily adopted or widely used by non-technical users, which constitute the bulk of the stakeholders. Importantly, cost-sensitive analysis, such as in this work, could perhaps inform new hybrid CFAs that may play a role in dementia diagnosis. Notably, our identified optimal set of assessment items included several items from the Functional Assessment Questionnaire (FAQ), which is often used in research study settings but less so in clinical practice. The effectiveness and high predictiveness of FAQ for AD diagnosis has also been identified in our previous work and the work of others [24], [5], [20].

Our analysis here is contingent on our estimation of the time duration to conduct an assessment item. Individuals with clinical expertise in performing these assessments might estimate assessment times differently from our estimates e.g. depending on their experience. This has motivated our development of a GUI-based sandbox-like tool which enables non-technical users to assign cost values (assessment time) to any of the CFA assessment items and specify parameters to perform cost-sensitive feature selection on the dataset. In the spirit of machine learning model operationalisation management (MLOps) [47], [48], we hope our GUI tool can help bridge the gap between machine learning research and other communities and stakeholders, especially clinicians, health economists and policy-makers. This can be achieved by providing hands-on experience of analysing the effectiveness and efficiency of various diagnostic assessments. Moreover, the intuitive use of our GUI tool can guide pedagogical training in health, biomedical or data science courses on cost-sensitive optimisation. However, it should be noted that clinical diagnosis involves assessments beyond CFAs, and that consideration of dementia assessment redesigning will have to involve other resource constraints e.g. financial costs of tests and biomarker acquisition [4], [49], [50]. These will be considered in future work.

To conclude, we have demonstrated the feasibility of cost-sensitive feature selection for Alzheimer’s disease diagnosis. The application of cost-sensitive feature selection within health and medical informatics is still an emerging field [16]. Given the importance of feature cost in a medical or clinical context, and the growing need to resolve overburdened healthcare systems, it is likely that this research area will continue to expand. In this work, although we have applied a time-based cost-sensitive feature selection algorithm to dementia CFAs, we foresee the same approach could be applied to many other medical and clinical settings where various types of costs or constraints have to be considered.

## Data Availability

The codes, including the RShiny code for the GUI, are available at: https://github.com/mac-n/Rshiny-app.

## Acknowledgment

This work was supported by the European Union’s INTERREG VA Programme, managed by the Special EU Programmes Body (SEUPB) (Centre for Personalised Medicine, IVA 5036)), and additional support by Alzheimer’s Research UK (ARUK) NI Pump Priming (XD, ST, PLM, KW-L) and Ulster University Research Challenge Fund (XD, ST, PLM, KW-L). The views and opinions expressed in this paper do not necessarily reflect those of the European Commission or the Special EU Programmes Body (SEUPB). We thank Daman Kaur for providing useful feedback on the work and the manuscript.

Data collection and sharing for this project was funded by the Alzheimer’s Disease Neuroimaging Initiative (ADNI) (National Institutes of Health Grant U01 AG024904) and DOD ADNI (Department of Defense award number W81XWH-12-2-0012). ADNI is funded by the National Institute on Aging, the National Institute of Biomedical Imaging and, and through generous contributions from the following: AbbVie, Alzheimer’s Association; Alzheimer’s Drug Discovery Foundation; Araclon Biotech; BioClinica, Inc.; Biogen; Bristol-Myers Squibb Company; CereSpir, Inc.; Cogstate; Eisai Inc.; Elan Pharmaceuticals, Inc.; Eli Lilly and Company; EuroImmun; F. Hoffmann-La Roche Ltd and its affiliated company Genentech, Inc.; Fujirebio; GE Healthcare; IXICO Ltd.; Janssen Alzheimer Immunotherapy Research Development, LLC.; Johnson Johnson Pharmaceutical Research Development LLC.; Lumosity; Lundbeck; Merck Co., Inc.; Meso Scale Diagnostics, LLC.; NeuroRx Research; Neurotrack Technologies; Novartis Pharmaceuticals Corporation; Pfizer Inc.; Piramal Imaging; Servier; Takeda Pharmaceutical Company; and Transition Therapeutics. The Canadian Institutes of Health Research is providing funds to support ADNI clinical sites in Canada. Private sector contributions are facilitated by the Foundation for the National Institutes of Health (www.fnih.org). The grantee organization is the Northern California Institute for Research and Education, and the study is coordinated by the Alzheimer’s Therapeutic Research Institute at the University of Southern California. ADNI data are disseminated by the Laboratory for Neuro Imaging at the University of Southern California.

