## Supplementary Materials for "Alzheimer’s Disease Assessments Optimised for Diagnostic Accuracy and Administration Time"

### Supplementary Algorithm 1

#### *Correlation-Based Feature Selection*[1], [2]

1. Set stopping criterion $n$ to avoid time-consuming search of the whole search space.
2. Initialise a best-first search through combinations of features with a start state – the empty set of features. Initialise an OPEN list containing the start state, a CLOSED list that is empty, and initialise the BEST combination as the start state.
3. Let $S$ be the set of features from OPEN with the highest merit. $Merit = \frac{k\overline{r_{cf}}}{\sqrt{k+k(k-1)\overline{r_{ff}}}}$. where $k$ is the number of features in S, $\overline{r_{cf}}$ is the mean feature-class correlation, and $\overline{r_{ff}}$ is the mean feature-feature correlation. The correlation between discrete features is measured by mutual information, and for continuous features the correlation coefficient is used.
4. Remove $S$ from OPEN and add to CLOSED
5. Generate all the children of $S$ by adding single features to $S$. Add each child of $S$ to the OPEN list.
6. If $Merit(S) > Merit(BEST)$, then set BEST to $S$.
7. If BEST changed in the last $n$ iterations, go to 2.
8. Return BEST.

### Supplementary Algorithm 2

#### *Consistency-Based Feature Selection*[2], [3]

1. Discretise the features.
2. Set stopping criterion $n$ to avoid time-consuming search of the whole search space.
3. Initialise a best-first search through combinations of features with a start state – the empty set of features. Initialise an OPEN list containing the start state, a CLOSED list that is empty, and initialise the BEST combination as the start state.
4. Let $S$ be the set of features from OPEN with the highest consistency measure. $Consistency$ is calculated using Liu's measure: number of **inconsistent examples/number of examples.** An "example" consists of the discretised features in $c$, plus the class value, of a single row from the dataset. An example is **inconsistent** where it has a different class value than the majority of examples with identical feature values.
5. Generate all the children of $S$ by adding single features to $S$. Add each child of $S$ to the OPEN list.
6. If $Consistency(\mathrm{S}) > Consistency(BEST)$ then set BEST to $S$
7. If BEST changed in the last $n$ iterations, go to 2.
8. Return BEST.

### Supplementary Algorithm 3

#### *Boruta Feature Selection*[4]

1. Duplicate the dataset to have two copies of each feature. The copied features are called shadow features.
2. Shuffle the values in each shadow feature randomly (shuffle the column). This means the shadow features carry no information about the class.
3. Build a Random Forest model on the dataset consisting of real and shadow features and extract the feature importance of all features.
4. All real features which outperform the highest ranked shadow feature score 1 point.
5. Repeat steps 1-4 20 times and generate the distribution of points for each (real) feature. Mark features as **selected** if they are in the highest tail of the distribution, **discarded** if they are in the lowest tail, and **tentative** if they are in between.

### Supplementary Figure 1

*Screenshot of R Shiny Application (available at https://mac-n.shinyapps.io/costcfs/)*

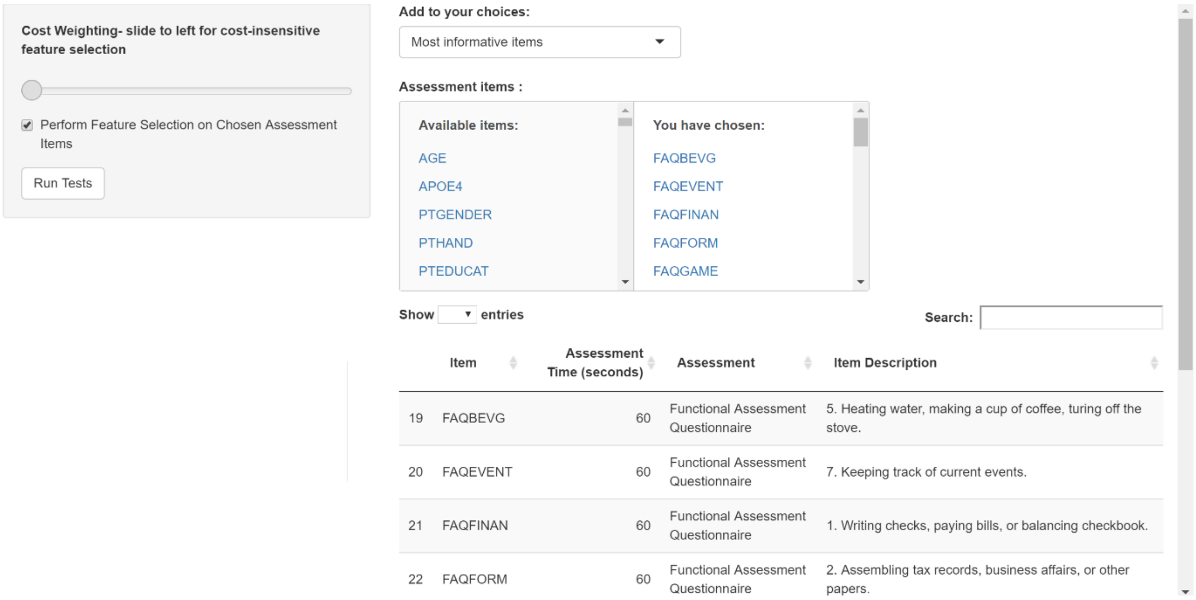

### Supplementary Table 1

#### *Details of all the cognitive and functional assessment (CFA) features used for feature selection*

| Feature | Source | Description |
|---|---|---|
| **AGE** | Demographics | Age |
| **PTEDUCAT** | Demographics | Education |
| **i6Serial** | MoCA[1] | Counting backwards |
| **visuo** | MoCA | Visual |
| **naming** | MoCA | Naming |
| **attention** | MoCA | Attention subscale |
| **language** | MoCA | Language subscale |
| **abstraction** | MoCA | abstraction Subscale |
| **delayedRecall** | MoCA | Delayed Word Recall |
| **orientation** | MoCA | Delayed Word Recall Subscale |
| **FAQFINAN** | FAQ[2] | 1. Writing checks, paying bills, or balancing checkbook. |
| **FAQFORM** | FAQ | 2. Assembling tax records, business affairs, or other papers. |
| **FAQSHOP** | FAQ | 3. Shopping alone for clothes, household necessities, or groceries. |
| **FAQGAME** | FAQ | 4. Playing a game of skill such as bridge or chess, working on a hobby. |
| **FAQBEVG** | FAQ | 5. Heating water, making a cup of coffee, turing off the stove. |
| **FAQMEAL** | FAQ | 6. Preparing a balanced meal. |
| **FAQEVENT** | FAQ | 7. Keeping track of current events. |
| **FAQTV** | FAQ | 8. Paying attention to and understanding a TV program, book, or magazine. |
| **FAQREM** | FAQ | 9. Remembering appointments, family occasions, holidays, medications. |
| **FAQTRAVL** | FAQ | 10. Traveling out of the neighborhood, driving, or arranging to take public transportation. |
| **MMDATE** | MMSE[3][4] | 1. What is today's date? |
| **MMYEAR** | MMSE | 2. What is the year? |
| **MMMONTH** | MMSE | 3. What is the month? |
| **MMDAY** | MMSE | 4. What day of the week is today? |
| **MMSEASON** | MMSE | 5. What season is it? |
| **MMHOSPIT** | MMSE | 6. What is the name of this hospital (clinic, place)? |
| **MMFLOOR** | MMSE | 7. What floor are we on? |
| **MMCITY** | MMSE | 8. What town or city are we in? |
| **MMAREA** | MMSE | 9. What county (district, borough, area) are we in? |
| **MMSTATE** | MMSE | 10. What state are we in? |
| **MMobjects** | MMSE | Composite: MMBALL,MMTREE,MMFLAG, MMBALLDL,MMTREEDL,MMFLAGDL. Identify 3 pictured objects. Name them again after a delay. |
| **MMWORLD** | MMSE | Composite: MMW,MMO,MMR,MML,MMD. “Spell WORLD backwards.” |
| **MMWATCH** | MMSE | 22. Show the participant a wrist watch and ask "What is this?" |

[1] Montreal Cognitive Assessment
[2] Functional Assessment Questionnaire
[3] Mini Mental State Examination
[4] Where some items from an assessment scale are not included, this is due to the quantity of missing data

| Feature | Source | Description |
| --- | --- | --- |
| **MMPENCIL** | MMSE | 23. Repeat for pencil. |
| **MMREPEAT** | MMSE | 24. Say, "Repeat after me: no ifs, ands, or buts." |
| **MMHAND** | MMSE | 25. Takes paper in right hand. |
| **MMFOLD** | MMSE | 26. Folds paper in half. |
| **MMONFLR** | MMSE | 27. Puts paper on floor. |
| **MMREAD** | MMSE | 28. Present the piece of paper which reads, "CLOSE YOUR EYES," and say: "Read this and do what it says." |
| **MMWRITE** | MMSE | 29. Give the participant a blank piece of paper and say: "Write a sentence." |
| **MMDRAW** | MMSE | 30. Present the participant with the Construction Stimulus page. Say, "Copy this design." |
| **Q1SCORE** | ADAS[5] | Word Recall Task |
| **Q2SCORE** | ADAS | Commands (receptive speech) |
| **Q3SCORE** | ADAS | Constructional Praxis (geometric figures) |
| **Q4SCORE** | ADAS | Delayed Word Recall |
| **Q5SCORE** | ADAS | Naming Task |
| **Q6SCORE** | ADAS | Ideational Praxis (sequence of familiar actions) |
| **Q7SCORE** | ADAS | Orientation (date/place etc) |
| **Q8SCORE** | ADAS | Word Recognition |
| **Q9SCORE** | ADAS | Remembering Test Instructions |
| **Q10SCORE** | ADAS | Comprehension (of examiner's speech) |
| **Q11SCORE** | ADAS | Word Finding Difficulty (in subject's speech) |
| **Q12SCORE** | ADAS | Spoke Language Ability (clarity of communication) |
| **Q13SCORE** | ADAS | Number Cancellation |
| **RAVLT.immediate** | RAVLT[6] | Rey Auditory Verbal Learning Test Total |
| **AVDELTOT** | RAVLT- Delayed | 30 Minute Delay Total |
| **CATANIMSC** | Category Fluency | Category Fluency (Animals) - Total Correct |
| **BNTTOTAL** | Boston Naming Test | Boston Naming Test Total Score |
| **CLOCKSCOR** | Clock Drawing | Total Score |
| **COPYSCOR** | Clock Copying | Total Score |
| **TRAASCOR** | Trail Making | Part A - Time to Complete |
| **TRABSCOR** | Trail Making | Trails B |
| **GDSATIS** | GDS[7] | 1. Are you basically satisfied with your life? |
| **GDDROP** | GDS | 2. Have you dropped many of your activities and interests? |
| **GDEMPTY** | GDS | 3. Do you feel that your life is empty? |
| **GDBORED** | GDS | 4. Do you often get bored? |
| **GDSPIRIT** | GDS | 5. Are you in good spirits most of the time? |
| **GDAFRAID** | GDS | 6. Are you afraid that something bad is going to happen to you? |
| **GDHAPPY** | GDS | 7. Do you feel happy most of the time? |
| **GDHELP** | GDS | 8. Do you often feel helpless? |
| **GDHOME** | GDS | 9. Do you prefer to stay at home, rather than going out and doing new things? |
| **GDMEMORY** | GDS | 10. Do you feel you have more problems with memory than most? |

[5] Alzheimer's Disease Assessment Scale
[6] Rey Auditory Verbal Learning Test
[7] Geriatric Depression Scale

| Feature | Source | Description |
|---|---|---|
| **GDALIVE** | GDS | 11. Do you think its wonderful to be alive now? |
| **GDWORTH** | GDS | 12. Do you feel pretty worthless the way you are now? |
| **GDENERGY** | GDS | 13. Do you feel full of energy? |
| **GDHOPE** | GDS | 14. Do you feel that your situation is hopeless? |
| **GDBETTER** | GDS | 15. Do you think that most people are better off than you are? |
| **DIVATT1** | ECog[8] - Patient | 1. The ability to do two things at once. |
| **DIVATT2** | ECog - Patient | 2. Returning to a task after being interrupted. |
| **DIVATT3** | ECog - Patient | 3. The ability to concentrate on a task without being distracted by external things in the environment. |
| **DIVATT4** | ECog - Patient | 4. Cooking or working and talking at the same time. |
| **LANG1** | ECog - Patient | 1. Forgetting the names of objects. |
| **LANG2** | ECog - Patient | 2. Verbally giving instructions to others. |
| **LANG3** | ECog - Patient | 3. Finding the right words to use in conversations. |
| **LANG4** | ECog - Patient | 4. Communicating thoughts in a conversation. |
| **LANG5** | ECog - Patient | 5. Following a story in a book or on TV. |
| **LANG6** | ECog - Patient | 6. Understanding the point of what other people are trying to say. |
| **LANG7** | ECog - Patient | 7. Remembering the meaning of common words. |
| **LANG8** | ECog - Patient | 8. Describing a program I have watched on TV. |
| **LANG9** | ECog - Patient | 9. Understanding spoken directions or instructions. |
| **MEMORY1** | ECog - Patient | 1. Remembering a few shopping items without a list. |
| **MEMORY2** | ECog - Patient | 2. Remembering things that happened recently (such as recent outings, events in the news). |
| **MEMORY3** | ECog - Patient | 3. Recalling conversations a few days later. |
| **MEMORY4** | ECog - Patient | 4. Remembering where I have placed objects. |
| **MEMORY5** | ECog - Patient | 5. Repeating stories and/or questions. |
| **MEMORY6** | ECog - Patient | 6. Remembering the current date or day of the week. |
| **MEMORY7** | ECog - Patient | 7. Remembering I have already told someone something. |
| **MEMORY8** | ECog - Patient | 8. Remembering appointments, meetings, or engagements. |
| **ORGAN1** | ECog - Patient | 1. Keeping living and work space organized. |
| **ORGAN2** | ECog - Patient | 2. Balancing the checkbook without error. |
| **ORGAN3** | ECog - Patient | 3. Keeping financial records organized. |
| **ORGAN4** | ECog - Patient | 4. Prioritizing tasks by importance. |
| **ORGAN5** | ECog - Patient | 5. Keeping mail and papers organized. |
| **ORGAN6** | ECog - Patient | 6. Using an organized strategy to manage a medication schedule involving multiple medications. |
| **PLAN1** | ECog - Patient | 1. Planning a sequence of stops on a shopping trip. |
| **PLAN2** | ECog - Patient | 2. The ability to anticipate weather changes and plan accordingly (i.e., bring a coat or umbrella) |
| **PLAN3** | ECog - Patient | 3. Developing a schedule in advance of anticipated events. |
| **PLAN4** | ECog - Patient | 4. Thinking things through before acting. |
| **PLAN5** | ECog - Patient | 5. Thinking ahead. |
| **VISSPAT1** | ECog - Patient | 1. Following a map to find a new location. |
| **VISSPAT2** | ECog - Patient | 2. Reading a map and helping with directions when someone else is driving. |

[8] Everyday Cognition

| Feature | Source | Description |
|---|---|---|
| **VISSPAT3** | ECog - Patient | 3. Finding my car in a parking lot. |
| **VISSPAT4** | ECog - Patient | 4. Finding my way back to a meeting spot in the mall or other location. |
| **VISSPAT6** | ECog - Patient | 5. Finding my way around a familiar neighborhood. |
| **VISSPAT7** | ECog - Patient | 6. Finding my way around a familiar store. |

#### Supplementary Table 2

*All cognitive and functional assessment (CFA) features selected more than once by feature selection algorithms*

| Name | # times selected | Estimated time (sec) | Notes on time estimation |
|---|---|---|---|
| faq_FAQSHOP | 14 | 60 | FAQ - total time 10 minutes, 10 items [5] |
| faq_FAQREM | 13 | 60 | FAQ - total time 10 minutes, 10 items [5] |
| faq_FAQTRAVL | 13 | 60 | FAQ - total time 10 minutes, 10 items [5] |
| adas_Q4SCORE | 11 | 600 | ADAS – total time 30-40 minutes [5], 13 items but 9,10,11 are not standalone assessment items. Q4 is delayed word recall - can be included in an assessment if Q1 is also included. Time estimation includes delay. |
| faq_FAQEVENT | 11 | 60 | FAQ - total time 10 minutes, 10 items [5] |
| faq_FAQFINAN | 11 | 60 | FAQ - total time 10 minutes, 10 items [5] |
| faq_FAQFORM | 11 | 60 | FAQ - total time 10 minutes, 10 items [5] |
| gd_GDMEMORY | 11 | 28 | Geriatric Depression Scale[5]: 5-7 minutes, 15 items |
| faq_FAQMEAL | 10 | 60 | FAQ - total time 10 minutes, 10 items [5] |
| RAVLT.immediate | 10 | 900 | total time for RAVLT 10-15 minutes[6] |
| adas_Q1SCORE | 9 | 200 | ADAS – total time 30-40 minutes [5], 13 items but 9,10,11 are not standalone assessment items. |
| BNTTOTAL | 9 | 600 | Boston Naming Test. 60 item version takes 10-20 minutes [6]. This is the 30 item version. |
| CATANIMSC | 9 | 150 | estimation from ADNI procedures manual (adni.loni.usc.edu). Instructions, 20 second trial, further instructions, one minute for the task. |
| ecog_MEMORY1 | 9 | 60 | Ecog - no total time available, forms mailed in advance for study ,estimated each at 60 based on FAQ |
| adas_Q7SCORE | 8 | 200 | ADAS – total time 30-40 minutes [5], 13 items but 9,10,11 are not standalone assessment items. |
| faq_FAQGAME | 8 | 60 | FAQ - total time 10 minutes, 10 items [5] |
| faq_FAQTV | 8 | 60 | FAQ - total time 10 minutes, 10 items [5] |
| AVDELTOT | 7 | 2000 | This is delayed RAVLT. 30 minute delay included in estimation |
| ecog_LANG9 | 7 | 60 | Ecog - no total time available, forms mailed in advance for study ,estimated each at 60 based on FAQ |
| TRABSCOR | 7 | 600 | Trail Making Test part B. Total time for parts A and B 5-10 minutes [6]. Part B builds on part A. |
| mmse_objects | 6 | 300 | Composite of MMBALL, MMTREE,MMSE, MMFLAG, MMBALLDL,MMFLAGDL,MMTREEDL. 20 seconds per item. 3 minute delay as referred to in ADNI procedures manual. |
| adas_Q8SCORE | 5 | 200 | ADAS – total time 30-40 minutes [5], 13 items but 9,10,11 are not standalone assessment items. |
| ecog_PLAN3 | 5 | 60 | Ecog - no total time available, forms mailed in advance for study ,estimated each at 60 based on FAQ |
| ecog_LANG2 | 4 | 60 | Ecog - no total time available, forms mailed in advance |

| Name | # times selected | Estimated time (sec) | Notes on time estimation |
|---|---|---|---|
| | | | for study ,estimated each at 60 based on FAQ |
| ecog_PLAN4 | 4 | 60 | Ecog - no total time available, forms mailed in advance for study ,estimated each at 60 based on FAQ |
| faq_FAQBEVG | 4 | 60 | FAQ - total time 10 minutes, 10 items [5]. |
| mmse_MMDATE | 4 | 20 | MMSE: 10 minutes [6] 30 point scale, 20 seconds per point. |
| ecog_LANG7 | 3 | 60 | Ecog - no total time available, forms mailed in advance for study ,estimated each at 60 based on FAQ |
| ecog_MEMORY2 | 3 | 60 | Ecog - no total time available, forms mailed in advance for study ,estimated each at 60 based on FAQ |
| ecog_MEMORY3 | 3 | 60 | Ecog - no total time available, forms mailed in advance for study ,estimated each at 60 based on FAQ |
| gd_GDENERGY | 3 | 28 | Geriatric Depression Scale [5]: 5-7 minutes, 15 items |
| mmse_MMDRAW | 3 | 40 | MMSE: 10 minutes [6] usually 20 seconds per point. This is the "draw a pentagon" item and 20 seconds is not realistic. |
| mmse_MMSEASON | 3 | 20 | MMSE: 10 minutes [6] 30 point scale, 20 seconds per point. |
| adas_Q10SCORE | 2 | 1800 | ADAS – total time 30-40 minutes [5], this is not a standalone assessment item but assessed throughout the entire ADAS assessment. |
| ecog_DIVATT4 | 2 | 60 | Ecog - no total time available, forms mailed in advance for study ,estimated each at 60 based on FAQ |
| ecog_MEMORY4 | 2 | 60 | Ecog - no total time available, forms mailed in advance for study ,estimated each at 60 based on FAQ |
| ecog_ORGAN6 | 2 | 60 | Ecog - no total time available, forms mailed in advance for study ,estimated each at 60 based on FAQ |
